# Prognostic Value of Coronary CT in Outpatients with Chest Pain: A 5-Year MACE Analysis Stratified by ASCVD Risk

**DOI:** 10.1101/2025.10.01.25337132

**Authors:** Dongwon Kim, Jinha Lee, Hyunhee Choi, Taesun Kim, Minjung Kim, Agnes S. Kim, Michael Azrin, Juyong Lee, Kyungsoon Hong, Kyutae Park

**Affiliations:** Cardiovascular Center, Chuncheon Sacred Heart Hospital, Hallym University College of Medicine, Chuncheon, Gangwon-do, Republic of Korea; Calhoun Cardiology Center, School of Medicine, University of Connecticut, Farmington CT, USA; Center for Population Health, UConn Health, Farmington, CT, USA

**Keywords:** ASCVD score, CCTA, MACE, coronary artery disease, stable chest pain

## Abstract

**Background:** Coronary computed tomography angiography (CCTA) is recommended for the diagnosis of initial coronary artery disease (CAD) in patients with stable chest pain. We sought to understand the prevalence and severity of coronary stenosis observed via CCTA and to determine how integrating these anatomical findings with conventional 10-year atherosclerotic cardiovascular disease (ASCVD) scores could enhance risk stratification and guide clinical decisions.

**Methods:** This was an open-label, prospective, single-center observational study including 1,492 outpatients with stable chest pain who underwent CCTA. We collected data on ASCVD risk factors and followed up patients for 5 years to monitor for major adverse cardiovascular events (MACE). We analyzed the prevalence of obstructive CAD (OCAD, ≥50% stenosis) across different ASCVD risk categories and its relationship with MACE.

**Results:** Among 1,492 patients, CCTA revealed OCAD in 16.0%. Over a 5-year follow-up, 7.2% of patients experienced MACE. The presence of OCAD significantly improved MACE prediction beyond ASCVD scores alone. Notably, patients with ASCVD < 7.5% and OCAD had a significantly higher MACE risk (adjusted hazards ratio: 3.634; p = 0.023) compared with those without OCAD. The highest risk was found in the ASCVD ≥ 7.5% with OCAD group (adjusted hazards ratio: 5.101; p<0.001).

**Conclusions:** CCTA provides significant incremental value for risk stratification in outpatients with stable chest pain. It helps uncover a high-risk group that might be underestimated by conventional ASCVD scores, supporting CCTA integration into clinical workups for earlier intervention and improved patient outcomes.

## Introduction

Over the last 50 years, the prevalence and mortality of cardiovascular disease (CVD) have been continuously rising; the associated socioeconomic costs have also increased^1^. Several studies have explored risk-evaluation tools for predicting CVD by identifying associated risk factors to prevent the increasing CVD mortality rate and guide clinical practice. After the establishment of the Diamond–Forrester (DF) classification, which briefly classifies angina based on symptoms^2^, the Framingham 10-year risk score (FRS), calculated by the Framingham coronary heart disease (CHD) prediction equation, was developed from a large-scale demographic study that identified several risk factors for CHD^3^. In 2013, the 10-year atherosclerotic CVD (ASCVD) risk score, calculated by the Pooled Cohort Risk Assessment Equations to estimate the 10-year ASCVD risk, was suggested in the American College of Cardiology/American Heart Association (ACC/AHA) guidelines^4^.

Chest pain is one of the most common reasons for outpatient cardiology evaluation, suggesting potential cardiac involvement^5–8^. However, in ambulatory settings with limited time and resources, it remains challenging to fully exclude significant coronary artery disease (CAD) based solely on symptoms or conventional risk scores. Despite the availability of tools such as the DF classification, FRS, and the 10-year ASCVD risk score, substantial evidence suggests that these models may underestimate risk in patients with subclinical disease, particularly when not accompanied by objective imaging^9,10^.

Coronary angiography is the most definitive test for confirming CAD; however, it includes an invasive procedure with associated risks. In addition, approximately only one-third of patients without a known disease who had undergone elective cardiac catheterization had obstructive CAD (OCAD) in the United States^11^. Therefore, noninvasive tests, such as exercise stress echocardiography, myocardial perfusion scan, coronary computed tomography angiography (CCTA), and cardiac magnetic resonance imaging, are expected to circumvent unnecessary coronary angiography and screen patients who require intervention. Among these assessments, CCTA has emerged as a reliable and noninvasive diagnostic modality with high negative predictive value and diagnostic accuracy for CAD^12–14^, which has been validated through landmark studies such as the Prospective Multicenter Imaging Study for Evaluation of Chest Pain (PROMISE) and Scottish Computer Tomography of the Heart Trial (SCOT-HEART). These trials demonstrated that CCTA not only improves diagnostic accuracy but also reduces unnecessary invasive testing, and may improve clinical outcomes in stable chest pain populations. The current European and American guidelines now support the use of CCTA as a first-line test in selected symptomatic patients, particularly when clinical risk stratification is inconclusive^5,17–19^.

Nevertheless, the role of CCTA in outpatient chest pain clinics—specifically as a complement or potential replacement to ASCVD risk-based assessments—has not been fully defined. The latest AHA/ACC guidelines for chest pain recommend no testing in patients with stable chest pain with no known CAD and low clinical risk assessment^5^. However, in real-world practice, many patients with low or borderline 10-year ASCVD risk scores may still harbor OCAD detectable only by imaging. Conversely, some high score patients may have no anatomic disease. This discordance raises critical questions about adequacy of the current risk assessment models and highlights the potential role of CCTA in improving individualized risk prediction.

Although traditional risk scores such as ASCVD predict long-term risk, their ability to identify near-term and event-prone patients remains limited. CCTA offers anatomical clarity that may enhance risk discrimination. This study aimed to evaluate whether adding CCTA findings, categorized by the Coronary Artery Disease-Reporting and Data System (CAD-RADS), improves prediction of 5-year major adverse cardiovascular events (MACE) beyond ASCVD risk alone.

This study is part of the CLEAR-OUT (Chest pain Longitudinal Evaluation with Advanced imaging for Risk stratification in an Outpatient cohort) initiative, a prospective registry that enrolled patients from a regional cardiology clinic in Jecheon, Korea. All participants underwent cardiac computed tomography (CT) as part of their initial diagnostic workup for stable chest pain and subsequently underwent a follow-up for up to 5 years to evaluate long-term cardiovascular outcomes. The CLEAR-OUT cohort was specifically designed to assess the real-world diagnostic yield and prognostic value of coronary CT for stratifying risk beyond traditional models such as the 10-year ASCVD risk score, with the ultimate goal of informing outpatient management strategies and reducing cardiovascular events through early intervention.

Our primary objective was to evaluate diagnostic and prognostic utility of CCTA in outpatients presenting with stable chest pain in a real-world setting, while the secondary objective was to determine whether adding CCTA findings could enhance risk stratification over ASCVD risk alone.

## Methods

### Study Population and Study Design

This was an open-label, prospective, single-center observational cohort study of outpatients presenting with stable chest pain who underwent CCTA between January 2017 and December 2020. Recognizing that traditional symptom-based classification and risk scores, such as 10-year ASCVD risk score, may not fully capture the underlying CAD, this study was designed to assess the diagnostic and prognostic utility of anatomic classification based on the CAD-RADS. We aimed to investigate how CCTA can enhance risk stratification and guide treatment in a real-world outpatient setting. This approach would allow us to objectively evaluate the presence and severity of coronary artery stenosis through CCTA, ultimately correlating these findings with 5-year MACE to provide real-world outcome data of patients with stable chest pain.

We included 1,492 patients in the final analysis out of 3,091 patients based on the following criteria: (1) age between 40 and 75 years, (2) availability of laboratory and clinical data to calculate the 10-year ASCVD risk score, (3) without previous history of ASCVD, and (4) a minimum of 6 months of outpatient follow-up after CT examination, with complete outcome data up to 5 years. All consecutive patients who met the eligibility criteria were recruited between January 2017 and December 2020. The patient inclusion and exclusion process is shown in Figure 1.

**Figure 1.**
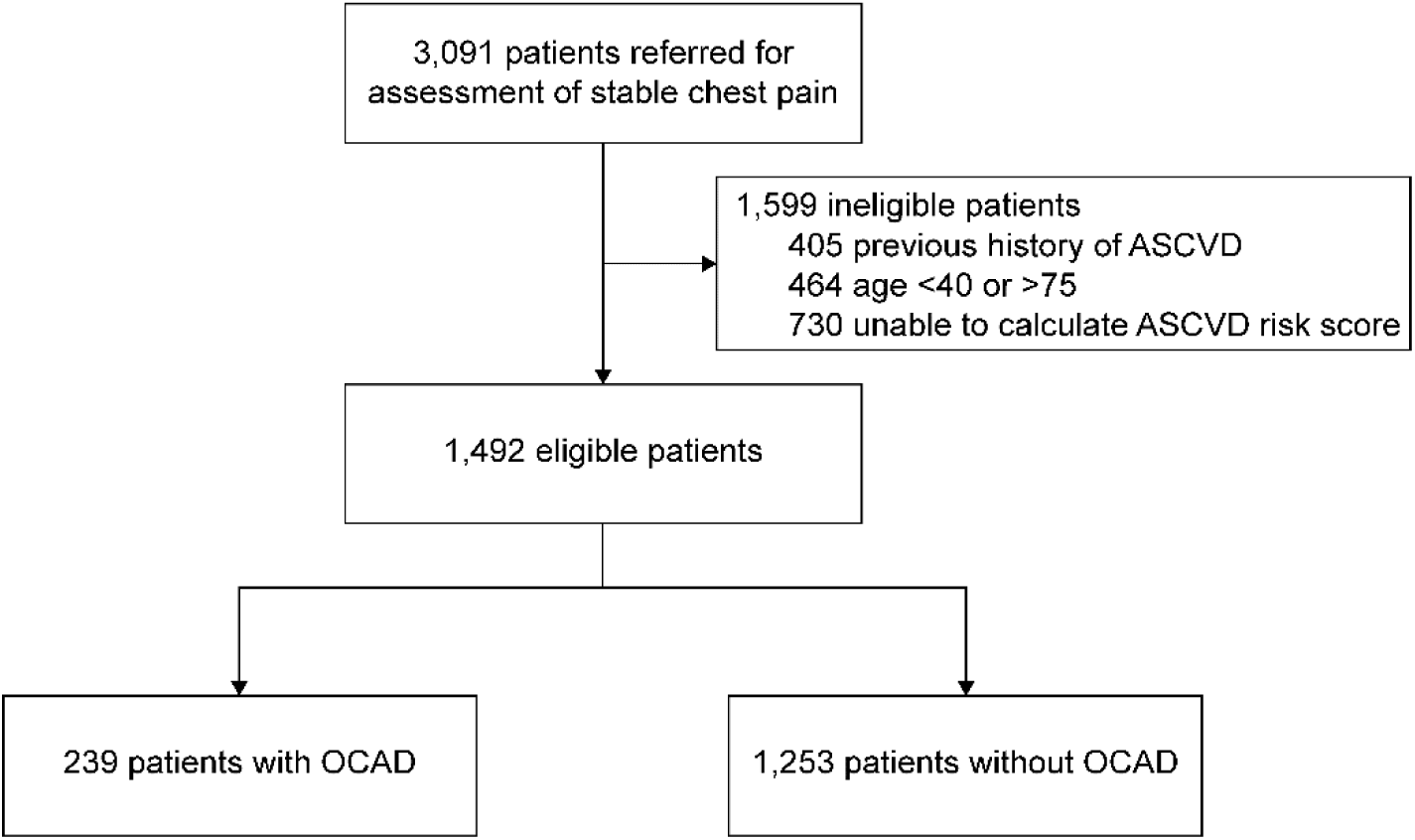
Flow diagram of patient selection ASCVD = atherosclerotic cardiovascular disease; OCAD = obstructive coronary artery disease

The study protocol was reviewed and approved by the relevant Institutional Review Board (IRB No. JCMJ 2020-01-001-001) and followed the principles of the latest version of the Declaration of Helsinki (2013). Written informed consent was obtained from all participants.

### Symptom Classification and Risk Stratification

For a direct comparison with CCTA findings, we classified patients using both symptom-based and traditional risk scores. The patients were classified as cardiac, possible cardiac, and noncardiac chest pain using DF classification and 2021 AHA/ACC chest pain guideline^2,5^. The 10-year ASCVD risk score was calculated by pooled cohort equations from the standard guidelines based on laboratory result at initial visit and stratified into four groups^3,4^:

- Low risk: <5%
- Borderline risk: 5.0–7.49%
- Intermediate risk: 7.5–19.99%
- High risk: ≥20%

For secondary analysis, ASCVD risk categories were dichotomized into two groups:

- ASCVD < 7.5%: The original low-to-borderline ASCVD risk groups
- ASCVD ≥ 7.5%: The original intermediate-to-high ASCVD risk groups

### Cardiac CT Protocol and Image Interpretation

All patients underwent non-contrast coronary artery calcium scoring (CACS), followed by CCTA using 64-slice multidetector CT scanner (GE Healthcare, Japan). Electrocardiography gating was used for image acquisition. To optimize image quality, the patients were premedicated to achieve a target heart rate of ≤70 beats per minute prior to CCTA. Ten mg oral propranolol was administered prior to hospital arrival in patients without contraindications. If the resting heart rate remained above target at the time of scanning, an additional 10 mg oral dose was considered. In patients exhibiting significant pre-procedural anxiety or discomfort, 2 mg oral diazepam was selectively administered to promote relaxation and assist in heart rate reduction. All medications were administered under appropriate clinical monitoring, and the decision to administer additional medication was based on resting heart rate, blood pressure, and patient symptoms at the time of evaluation.

CACS was categorized into four groups: 0, 1–99, 100–299, and ≥300. In addition, coronary stenosis severity was stratified according to the CAD-RADS classification16^)17^:

- CAD-RADS 0: 0% stenosis (no plaque or stenosis)
- CAD-RADS 1: 1–24% stenosis (minimal stenosis or plaque without stenosis)
- CAD-RADS 2: 25–49% stenosis (mild stenosis)
- CAD-RADS 3: 50–69% stenosis (moderate stenosis)
- CAD-RADS 4A: 70–99% stenosis (severe stenosis)
- CAD-RADS 4B: Left main coronary artery > 50% stenosis or three vessel ≥70% stenosis
- CAD-RADS 5: 100% stenosis (Total occlusion)

OCAD was a key endpoint in our analysis and was defined as CAD-RADS 3-5, which meant ≥50% luminal narrowing in one or more coronary vessels, in line with the most recent ACC/AHA guidelines^5^. Multivessel disease was defined as ≥50% stenosis in ≥2 major epicardial arteries. In secondary analysis, CAD-RADS categories were dichotomized into four groups according to the recommended management consideration^17^:

- CAD-RADS 0 : 0% stenosis
- CAD-RADS 1-2 : 1-49% stenosis
- CAD-RADS 3 : 50-69% stenosis
- CAD-RADS 4-5 : 70-100% stenosis

### Follow-Up and Clinical Outcomes

All patients were followed up longitudinally for up to 5 years at least, with a median follow-up of 5.13 years. All event data were confirmed by chart review, visit records, and direct follow-up at outpatient clinic.

The primary outcome was the occurrence of MACE, defined as a composite of:

- Cardiovascular death
- Nonfatal myocardial infarction (MI)
- Nonfatal stroke
- Hospitalization for unstable angina (UA)
- Unplanned coronary revascularization (percutaneous coronary intervention (PCI) or Coronary Artery Bypass Grafting (CABG))

To mitigate the potential of CCTA findings for directly influencing clinicians’ decisions to perform revascularization procedures and artificially inflate MACE rates, leading to a biased interpretation of the study’s findings, we conducted a sensitivity analysis using an alternative endpoint, MACE excluding unplanned coronary revascularization (MACE-UCR).

### Statistical Analysis

Continuous variables were presented as mean ± standard deviation or median (interquartile range) and were compared using one-way analysis of analysis or Kruskal-Wallis test depending on their distribution. The normality of continuous variables including physical measurements and blood test results was assessed by visual inspection using histogram. Categorical variables, including sociodemographic characteristics, are expressed as percentages and ratios and were analyzed using chi-square tests or Fisher’s exact tests as appropriate. In addition, we constructed receiver operating characteristic (ROC) curves and calculated the area under the curve (AUC) to compare the diagnostic accuracy of the DF classification, FRS, 10-year ASCVD risk score, and CACS for predicting the presence of OCAD.

Cox proportional hazards models were used to evaluate the association of 5-year MACE with: (1) 10-year ASCVD risk groups alone, (2) CAD-RADS categories alone, and (3) the combined 10-year ASCVD risk groups and CCTA findings, computing hazard ratios (HR) and 95% confidence intervals (CI). The assumptions of the proportional-hazards model were evaluated by Schoenfeld residual. In our analysis, we initially observed that a standard Cox proportional hazards model for MACE across the entire study duration violated the proportional hazards assumption. Therefore, we conducted a landmark analysis at 3 months post-enrollment. This specific landmark time point was chosen because we noted a high frequency of events, such as coronary angiography or PCI, occurring within the initial 3 months of the study. By excluding patients who experienced MACE within this initial 3-month period, our landmark analysis allowed us to satisfy the proportional hazards assumption for events occurring after this point. This enabled us to conduct a valid and interpretable Cox regression analysis on the subsequent data, providing a more accurate understanding of long-term MACE risk. Kaplan-Meier survival curves and log-rank tests were employed to illustrate and compare the cumulative incidence of 5-year MACE by: (1) CAD-RADS categories alone, and (2) the combined 10-year ASCVD risk groups and CCTA findings.

All statistical analyses were performed using R software version 4.4.1 (R Foundation for Statistical Computing, Vienna, Austria). A two-sided P-value <0.05 was considered statistically significant. Missing values were not imputed; the number of missing values for each variable is presented in Table 1.

**Table 1.**
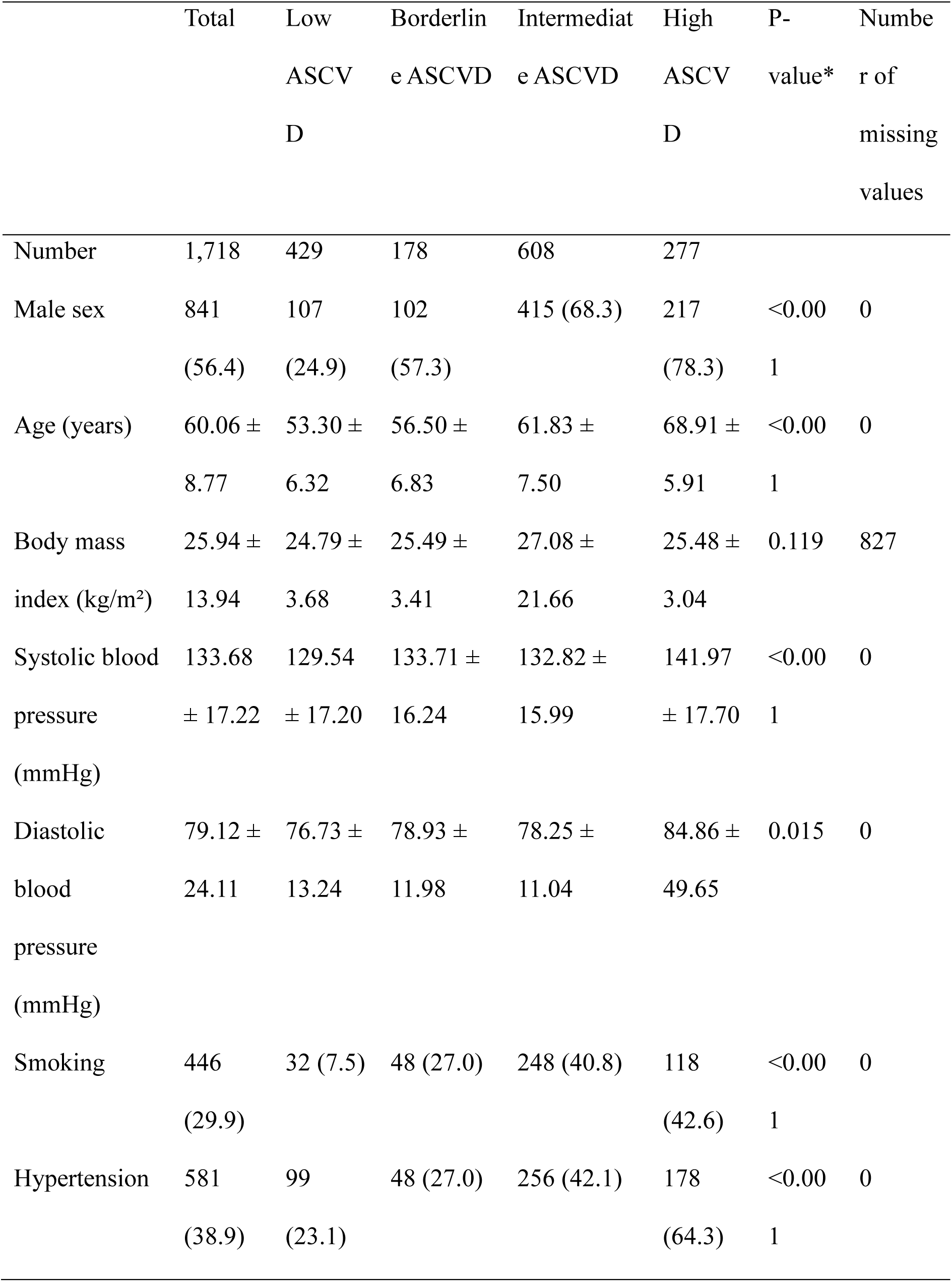
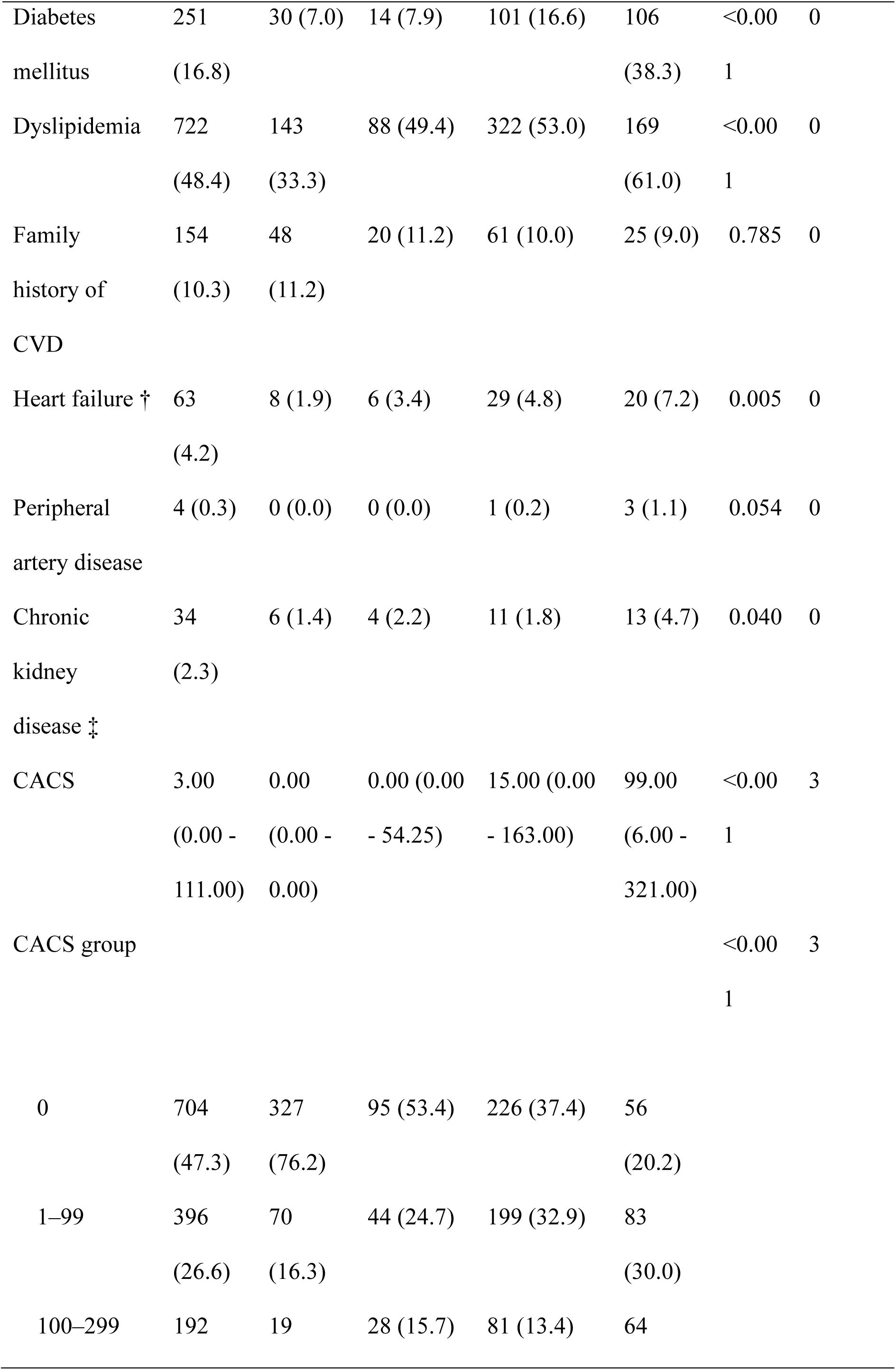
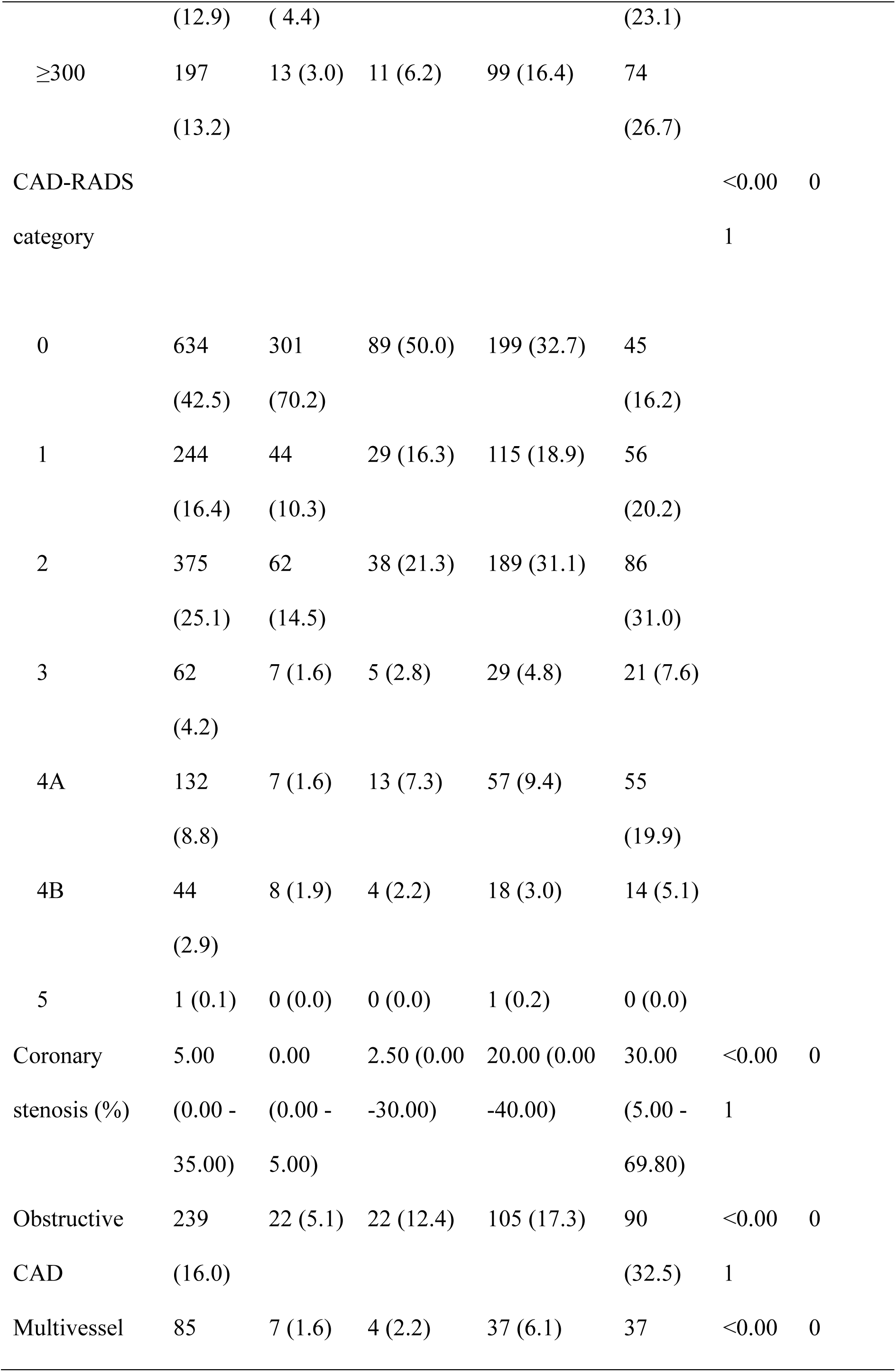
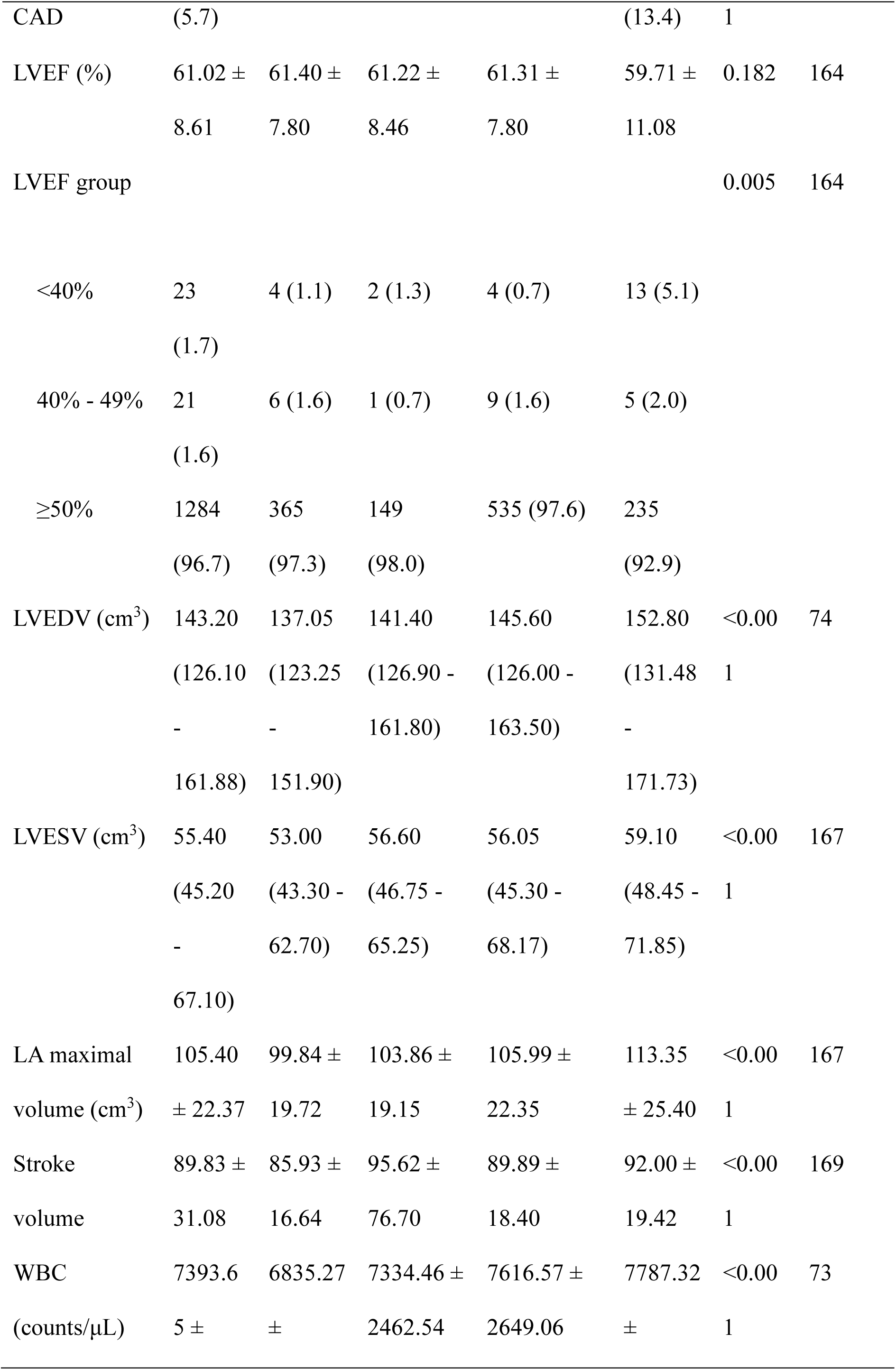
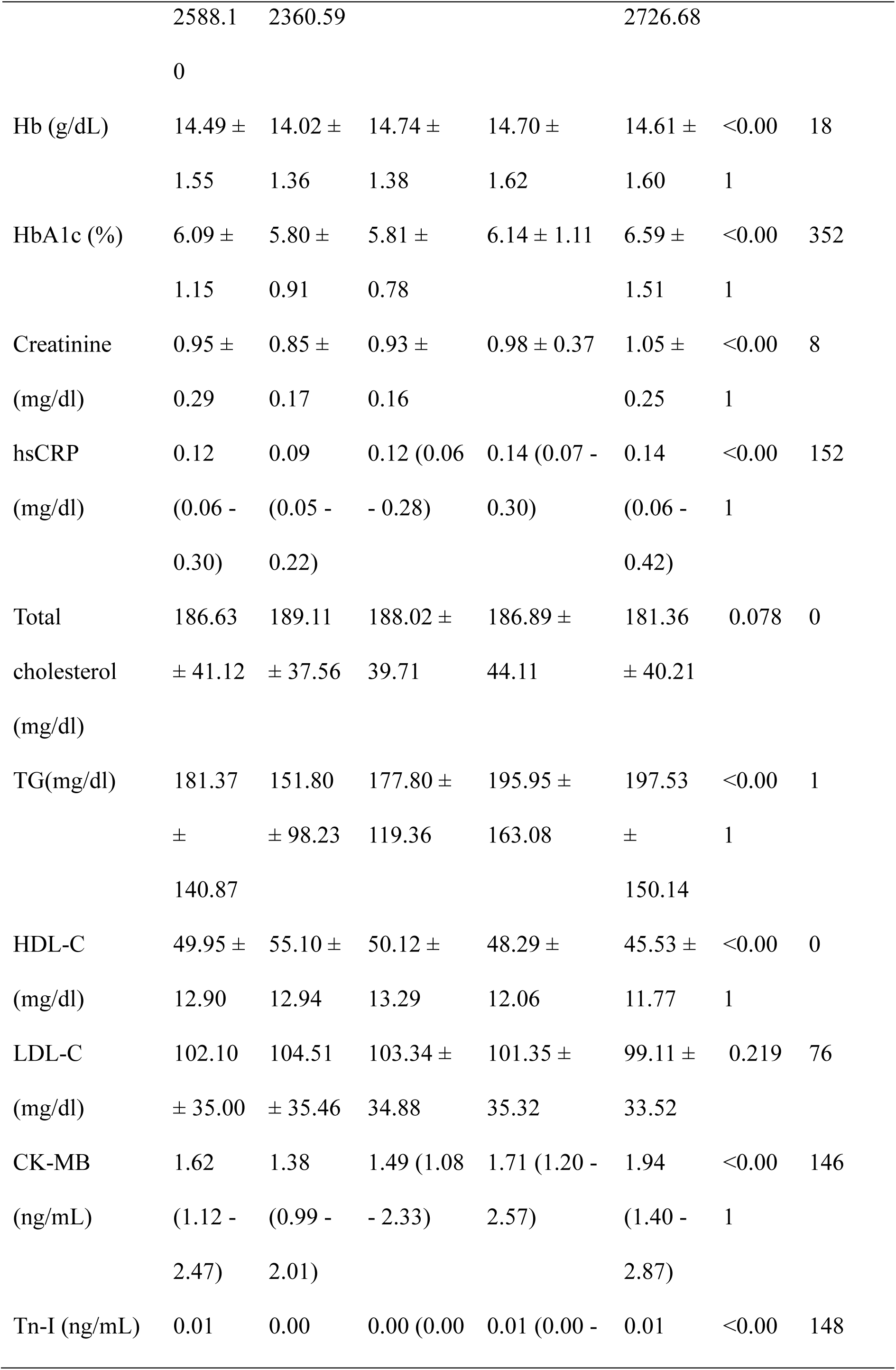
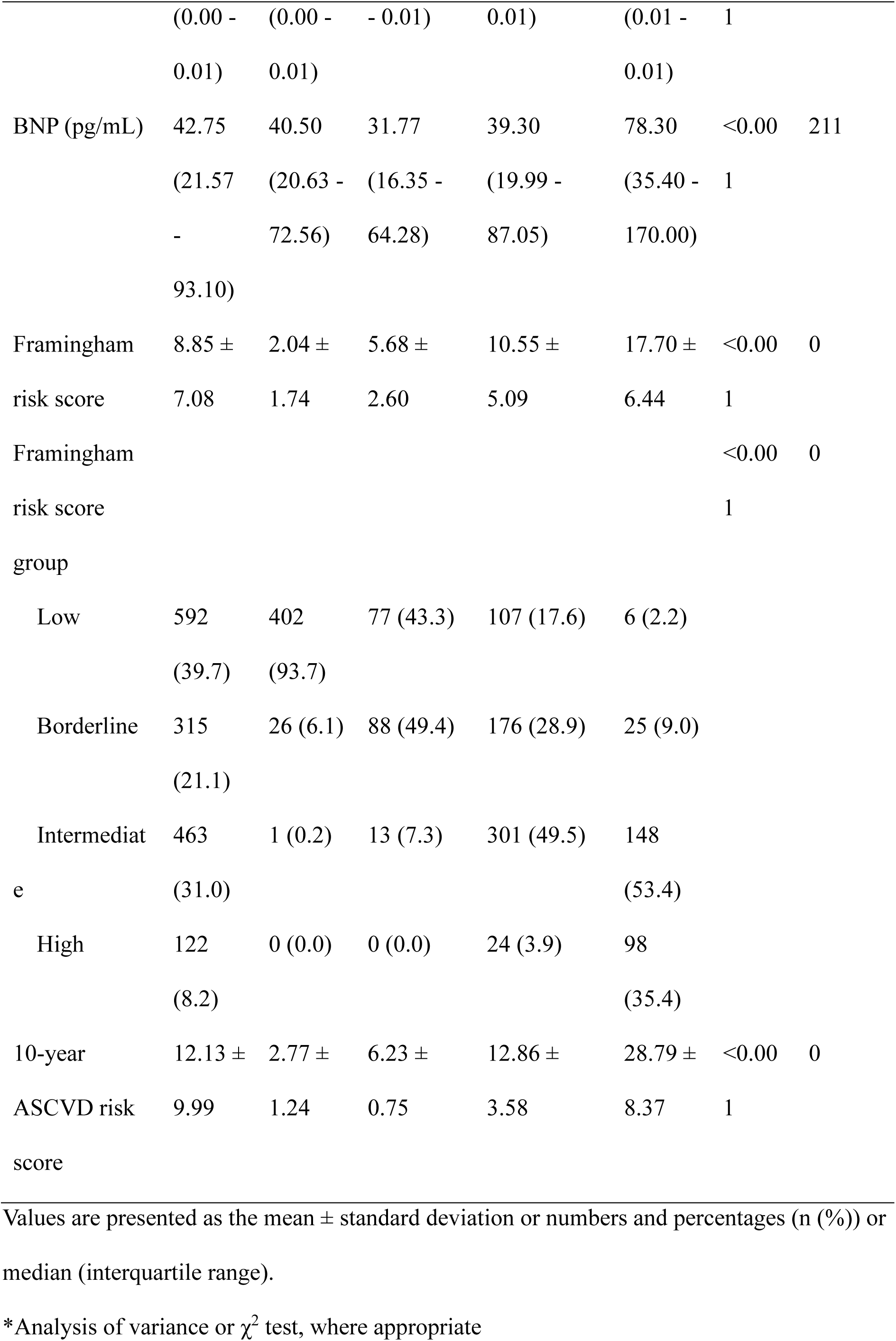
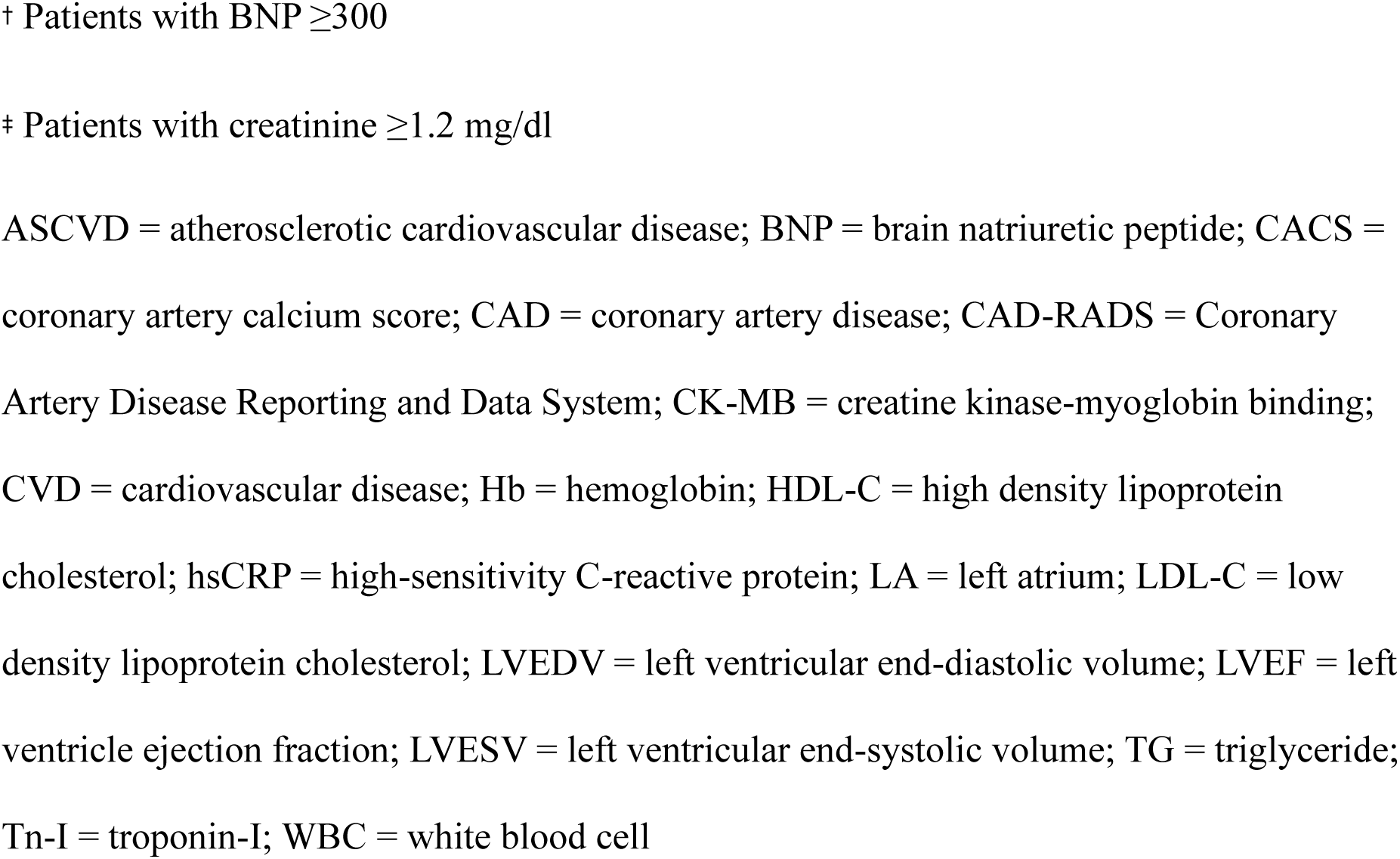
Baseline characteristics of the study population overall and across 10-year ASCVD risk groups.

## Results

### Baseline Characteristics

The baseline patient characteristics of 1,492 patients are presented in Table 1. The mean age was 60.06±8.77 years, and 841 (56.4%) were men. In total, 711 (47.7%) patients presented with noncardiac chest pain, while 275 (18.4%) and 506 (33.9%) presented with cardiac and possible cardiac chest pain, respectively. The mean FRS and 10-year ASCVD risk scores were 8.85 ± 7.08 and 12.13 ± 9.99, respectively. The median CACS was 3.00 (interquartile range: 0.00–111.00). Regarding other comorbidities, 581 (38.9%) patients reported having hypertension, 251 (16.8%), diabetes mellitus (DM), and 173 (11.6%), dyslipidemia.

There was a clear trend of increasing traditional cardiovascular risk factors across higher ASCVD risk groups. Patients with higher groups of ASCVD risk were significantly older, more likely to be male, and had higher rates of smoking, hypertension, DM, dyslipidemia compared to those in lower risk groups. In contrast, there was no significant difference in low-density lipoprotein cholesterol across the ASCVD risk groups (p = 0.219). Importantly, on CCTA, patients with higher ASCVD risk groups were strongly associated with a greater burden of CAD, as evidenced by higher CACS, CAD-RADS category and more frequent presence of OCAD and multivessel CAD.

### Prevalence of OCAD

OCAD was present in 239 (16.0%) of all patients on CCTA. In contrast, 634 (42.5%) patients had no visible CAD (CAD-RADS 0). Sixty-two (4.2%), 132 (8.9%), 44 (3.0%), and one (0.1%) patients exhibited 50%–69% stenosis (CAD-RADS 3), 70%–99% stenosis (CAD-RADS 4a), ≥50% left main artery stenosis or ≥70% stenosis in three vessels (CAD-RADS 4b), and total occlusion (CAD-RADS 5), respectively.

In patients with ASCVD < 7.5%, 44 (7.3%) showed OCAD, while 195 (22.0%) in the ASCVD ≥ 7.5% group had OCAD. Despite an approximately three-fold significant difference in prevalence, OCAD was still clearly observed in the lower ASCVD group.

### Diagnostic Performance of Clinical Risk Scores for Detecting OCAD

The ROC curves and AUCs are shown in Figure 2. The AUC of the 10-year ASCVD risk score (AUC, 0.703; 95% CI, 0.668–0.739) was higher than that of the DF classification (AUC, 0.624; 95% CI, 0.585–0.663) and FRS (AUC, 0.657; 95% CI, 0.621–0.692), while the CACS showed the highest diagnostic accuracy, with an AUC of 0.814 (95% CI, 0.784–0.843). The AUC of the 10-year ASCVD risk score and CACS exhibited statistically significant difference compared with the AUC of the DF classification (p < 0.001), whereas the AUC of FRS was not significantly different from that of DF classification (p = 0.224).

**Figure 2.**
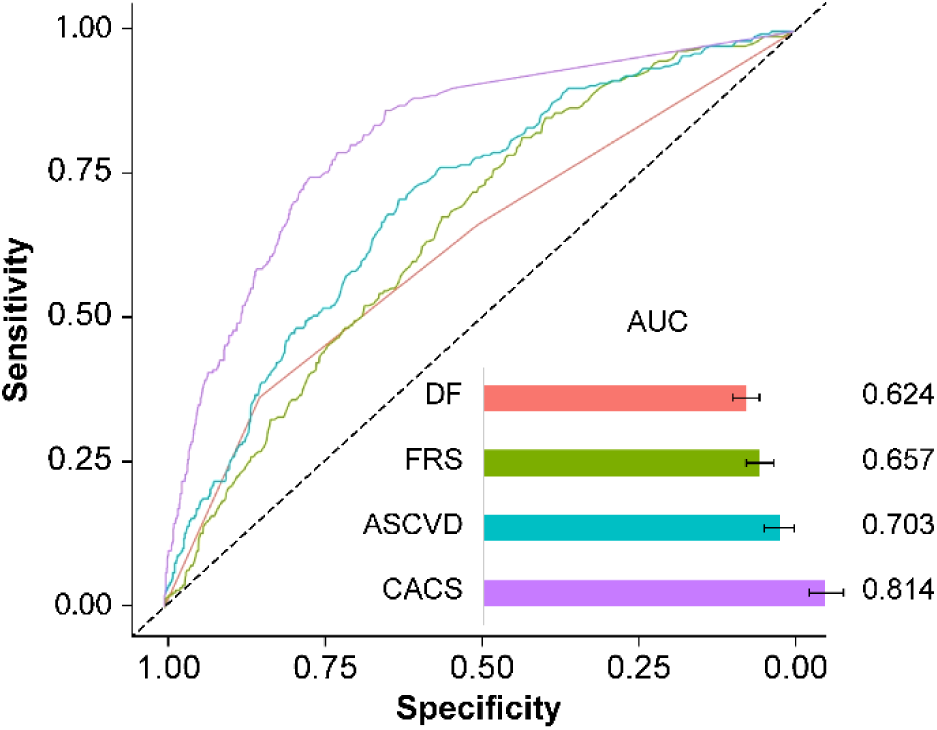
ROC curve and calculated AUCs of clinical risk scores in OCAD ROC Curves of DF classification, FRS, 10-year ASCVD risk score and CACS in the presence of OCAD are represented with their calculated AUCs, respectively. ASCVD = atherosclerotic cardiovascular disease; AUC = area under the curve; CACS = coronary artery calcium score; CAD = coronary artery disease; DF = Diamond-Forrester; FRS = Framingham risk score; OCAD = obstructive coronary artery disease; ROC = receiver-operating characteristic

### Association between Clinical Outcomes and ASCVD Risk

During the 5-year follow-up, a total of 108 patients (7.2%) experienced a MACE, including 17 (1.1%) cardiovascular death, 16 (1.1%) nonfatal MI, 11 (0.7%) nonfatal stroke, 48 (3.2%) hospitalization for UA, and 24 (1.6%) unplanned coronary revascularization. As summarized in Table 2, the cumulative incidence of 5-year MACE progressively increased from 3.3% in the low ASCVD group to 12.6% in the high ASCVD group (p < 0.001), consistent with traditional risk stratification. The patients in the higher ASCVD risk groups exhibited higher percentages of cardiovascular death (p = 0.040) and unplanned coronary revascularization (p < 0.001). However, there was no significant difference in the incidence of nonfatal MI (p = 0.474), nonfatal stroke (p = 0.355), and hospitalization for UA (p = 0.126) among these categories. In patients with ASCVD < 7.5%, 24 (4.0%) developed 5-year MACE, whereas 84 (9.5%) cases of ASCVD ≥ 7.5% had 5-year MACE. Although the 5-year MACE incidence in the ASCVD < 7.5% group was approximately half of the overall average, it is important to note that events still occurred in this group.

**Table 2.** Adverse cardiovascular events according to ASCVD risk groups during followup.

| Adverse cardiovascular events | Low ASCVD | Borderline ASCVD | Intermediate ASCVD | High ASCVD | P-value |
| --- | --- | --- | --- | --- | --- |
| 5-year MACE | 14 (3.3) | 10 (5.6) | 49 (8.1) | 35 (12.6) | <0.001 |
| Cardiovascular death | 2 (0.5) | 0 (0.0) | 8 (1.3) | 7 (2.5) | 0.037 |
| Nonfatal MI | 2 (0.5) | 2 (1.1) | 8 (1.3) | 4 (1.4) | 0.482 |
| Nonfatal stroke | 2 (0.5) | 0 (0.0) | 5 (0.8) | 4 (1.4) | 0.359 |
| Hospitalization for UA | 8 (1.9) | 5 (2.8) | 21 (3.5) | 14 (5.1) | 0.126 |
| Unplanned coronary revascularization | 0 (0.0) | 3 (1.7) | 10 (1.6) | 11 (4.0) | 0.001 |

Values are presented as the numbers and percentages (n (%)).
ASCVD = atherosclerotic cardiovascular disease; MACE = major adverse cardiac event; MI = myocardial infarction; UA = unstable angina

### Association between Clinical Outcomes and CCTA Findings Classified by CAD-RADS

Higher CAD-RADS categories were associated with a significantly greater percentage of 5-year MACE (p < 0.001), as listed in Table 3. The incidence of MACE was the lowest in the CAD-RADS 0 group (3.5%) and increased incrementally to 5.5% in the CAD-RADS 1-2 group, 21.0% in the CAD-RADS 3 group and was the highest in the CAD-RADS 4-5 group (22.0%) (p < 0.001). The patients in higher CAD-RADS categories also showed higher percentages of cardiovascular death (p = 0.017) and unplanned coronary revascularization (p < 0.001), whereas there was no significant difference in the incidence of nonfatal MI among these categories (p = 0.024).

**Table 3.** Adverse cardiovascular events according to CAD-RADS categories during.

| Adverse | CAD-RADS | CAD-RADS | CAD-RADS | CAD-RADS | P- |
| --- | --- | --- | --- | --- | --- |
| cardiovascular | 0 | 1-2 | 3 | 4-5 | value |
| events |  |  |  |  |  |
| 5-year MACE | 22 (3.5) | 34 (5.5) | 13 (21.0) | 39 (22.0) | <0.001 |
| Cardiovascular | 2 (0.3) | 9 (1.5) | 1 (1.6) | 5 (2.8) | 0.017 |
| death |  |  |  |  |  |
| Nonfatal MI | 2 (0.3) | 10 (1.6) | 0 (0.0) | 4 (2.3) | 0.024 |
| Nonfatal stroke | 5 (0.8) | 1 (0.2) | 0 (0.0) | 5 (2.8) | 0.009 |
| Hospitalization | 12 (1.9) | 13 (2.1) | 9 (14.5) | 14 (7.9) | <0.001 |
| for UA |  |  |  |  |  |
| Unplanned | 1 (0.2) | 3 (0.5) | 3 (4.8) | 17 (9.6) | <0.001 |
| coronary |  |  |  |  |  |
| revascularization |  |  |  |  |  |

Values are presented as the numbers and percentages (n (%)).
CAD-RADS = Coronary Artery Disease-Reporting and Data System; MACE = major adverse cardiac event; MI = myocardial infarction; OCAD = obstructive coronary artery disease; UA = unstable angina

The Kaplan-Meier survival analysis (Figure 3A) demonstrated a significant progressive increase in the cumulative incidence of 5-year MACE across the higher CAD-RADS categories (log-rank p < 0.001). The patients classified as CAD-RADS 0 exhibited the lowest cumulative incidence of MACE throughout the 5-year follow-up period. The cumulative incidence of MACE was progressively higher in patients with CAD-RADS 1-2, followed by CAD-RADS 3, and was the highest in the CAD-RADS 4-5 group. These findings confirm that CAD-RADS classification correlates with progressively increasing risk of MACE over 5 years. The sensitivity analysis using 5-year MACE-UCR yielded similar results (Figure S1A).

**Figure 3.**
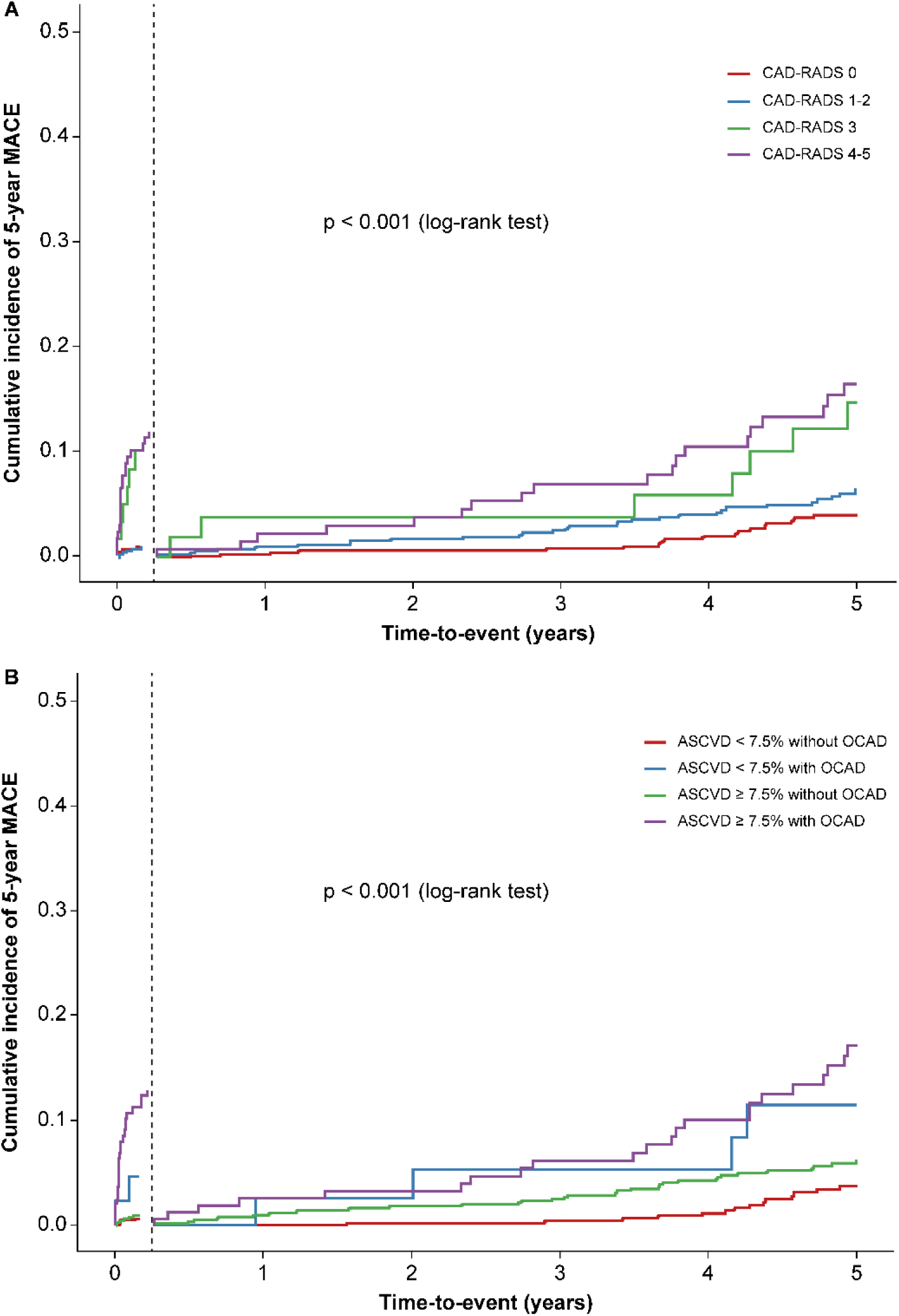
Cumulative incidence plot of the 5-year MACE stratified by (A) CAD-RADS categories (B) combined ASCVD risk groups and CCTA findings Kaplan-Meier survival analysis on 5-year MACE was conducted with landmark analysis 3 months after enrollment. ASCVD = atherosclerotic cardiovascular disease; CAD-RADS = Coronary Artery Disease-Reporting and Data System; MACE = major adverse cardiac event; OCAD = obstructive coronary artery disease

### Combined Prognostic Value of ASCVD and CCTA Findings

When we combined ASCVD risk groups with CCTA findings, the highest incidence of 5-year MACE was observed in the “ASCVD ≥ 7.5% with OCAD” group (23.6%), as presented in Table 4. Notably, the “ASCVD < 7.5% with OCAD” group had a MACE rate of 13.6%, which was more than double the rate of the “ASCVD ≥ 7.5% without OCAD” group (5.5%). This highlights the significant incremental value of CCTA findings in complementing and enhancing the prognostic accuracy of the traditional ASCVD risk score, particularly by identifying high-risk individuals underestimated by conventional models. The percentage of 5-year MACE, hospitalization for UA and unplanned coronary revascularization, was the highest in group with “ASCVD ≥ 7.5% with OCAD”, followed by “ASCVD < 7.5% with OCAD”, “ASCVD ≥ 7.5% without OCAD”, and “ASCVD < 7.5% without OCAD” (all, p < 0.001). However, there was no significant difference in the incidence of nonfatal MI among these categories (p = 0.240).

**Table 4.**
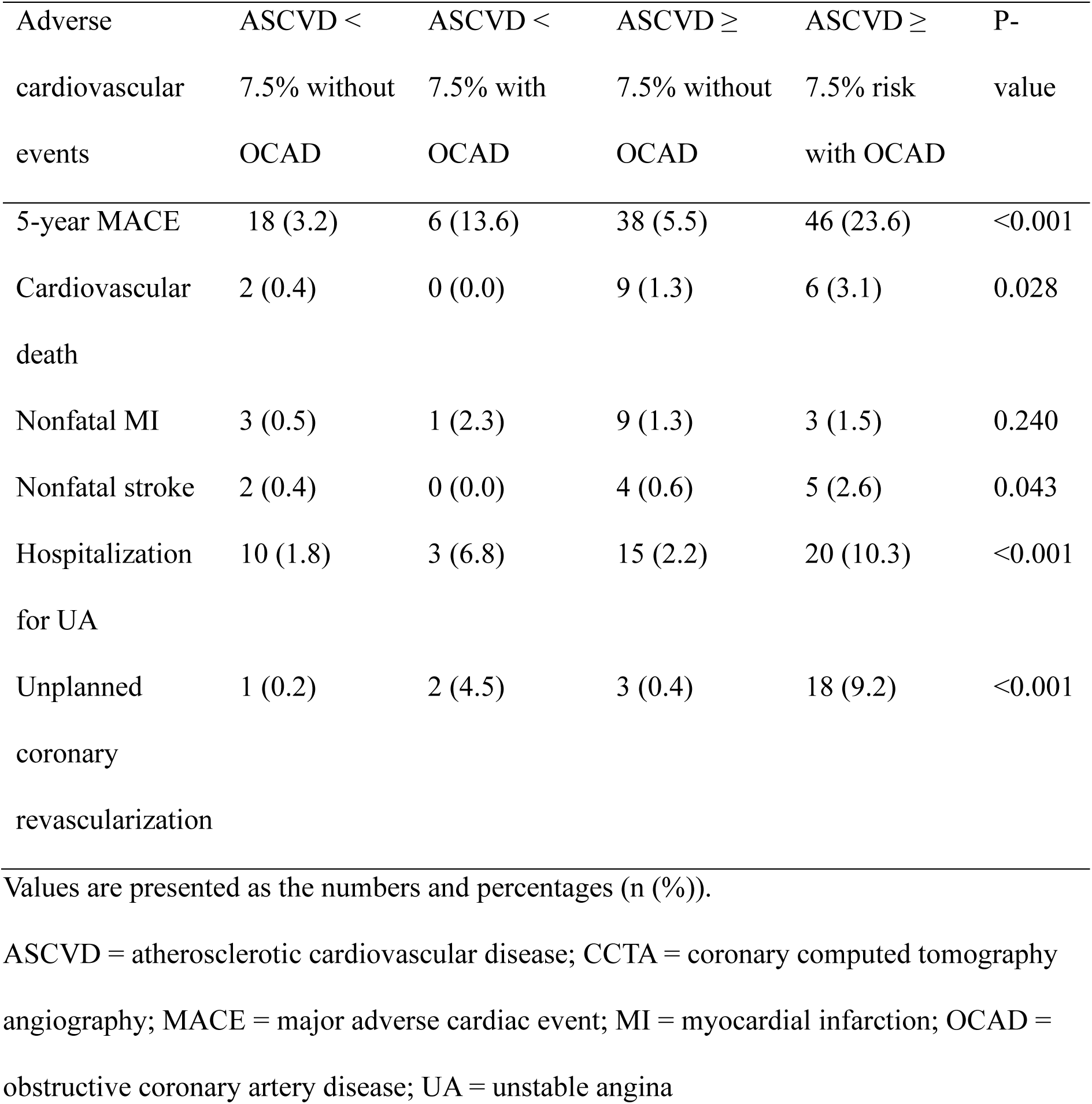
Adverse cardiovascular events according to combined ASCVD risk groups and CCTA findings during follow-up.

The Kaplan-Meier analysis (Figure 3B) showed the cumulative incidence of MACE to be the highest in the “ASCVD ≥ 7.5% with OCAD”, followed by “ASCVD < 7.5% with OCAD”, “ASCVD ≥ 7.5% without OCAD” and “ASCVD < 7.5% without OCAD” (log-rank p <0.001). The sensitivity analysis using 5-year MACE-UCR showed similar results (Figure S1B).

### Predictive Factors of MACE

Table 5 presents the univariate and multivariate Cox proportional hazard analyses of MACE according to ASCVD risk groups, CAD-RADS categories, and the combined ASCVD and CCTA findings. Regarding the multivariate analyses, adjustments were made for potential confounders including chronic kidney disease (CKD) and family history of CVD. Traditional risk factors such as age, sex, hypertension, DM, dyslipidemia, and smoking were excluded for adjustment as they are included in 10-year ASCVD risk calculation.

**Table 5.** Univariate and multivariate Cox proportional hazard analyses for the 5-year MACE according to the ASCVD risk groups, CAD-RADS categories, and combined ASCVD risk groups and CCTA findings

|  | Univariate analyses |  |  | Multivariate analyses* |  |  |
| --- | --- | --- | --- | --- | --- | --- |
|  | HR | 95% CI | P-value | aHR | 95% CI | P-value |
| ASCVD group |  |  |  |  |  |  |
| Low ASCVD | Reference |  |  | Reference |  |  |
| Borderline | 1.533 | 0.594- | 0.377 | 1.530 | 0.593- | 0.379 |
| ASCVD |  | 3.954 |  |  | 3.950 |  |
| Intermediate | 2.305 | 1.168- | 0.016 | 2.241 | 1.134- | 0.020 |
| ASCVD |  | 4.549 |  |  | 4.428 |  |
| High ASCVD | 2.972 | 1.424- | 0.004 | 2.800 | 1.335- | 0.006 |
|  |  | 6.203 |  |  | 5.873 |  |
| CAD-RADS category |  |  |  |  |  |  |
| CAD-RADS 0 | Reference |  |  | Reference |  |  |
| CAD-RADS 1-2 | 1.693 | 0.931- | 0.085 | 1.616 | 0.885- | 0.119 |
|  |  | 3.082 |  |  | 2.951 |  |
| CAD-RADS 3 | 4.026 | 1.669- | 0.002 | 3.897 | 1.610- | 0.003 |
|  |  | 9.709 |  |  | 9.434 |  |
| CAD-RADS 4-5 | 4.646 | 2.415- | <0.001 | 4.490 | 2.329- | <0.001 |
|  |  | 8.938 |  |  | 8.656 |  |
| ASCVD group+ CCTA finding |  |  |  |  |  |  |

| ASCVD <<br>7.5% without<br>OCAD | Reference |  |  | Reference |  |  |
| --- | --- | --- | --- | --- | --- | --- |
| ASCVD <<br>7.5% with<br>OCAD | 3.701 | 1.218-11.244 | 0.021 | 3.634 | 1.194-11.059 | 0.023 |
| ASCVD<br>≥7.5% without<br>OCAD | 1.921 | 1.025-3.600 | 0.041 | 1.840 | 0.979-3.458 | 0.058 |
| ASCVD<br>≥7.5% with<br>OCAD | 5.294 | 2.709-10.347 | <0.001 | 5.101 | 2.606-9.988 | <0.001 |

Cox proportional hazard analyses on 5-year MACE were performed with landmark analyses 3 months after enrollment.
\*Multivariate analyses were adjusted for CKD and family history of CVD. CKD is defined as creatinine $\geq 1.2$ mg/dl.
aHR = adjusted hazard ratio; ASCVD = atherosclerotic cardiovascular disease; CAD-RADS = Coronary Artery Disease-Reporting and Data System; CCTA = coronary computed tomography angiography; CI = confidence interval; CKD = chronic kidney disease; CVD = cardiovascular disease; HR = hazard ratio; hsCRP = high-sensitivity C-reactive protein; MACE = major adverse cardiac event; MI = myocardial infarction; OCAD = obstructive coronary artery disease; PCI = percutaneous coronary intervention

Considering the analysis based on the ASCVD risk groups alone, the Borderline, Intermediate, and High ASCVD groups showed progressively increasing HR for MACE compared with the Low ASCVD group. The Intermediate ASCVD group demonstrated a significantly higher risk using both univariate (HR, 2.305; 95% CI, 1.168–4.549; p = 0.016) and multivariate analyses (adjusted HR [aHR], 2.241; 95% CI, 1.134–4.428; p = 0.020). The High ASCVD group exhibited the highest risk, with HRs of 2.972 (95% CI, 1.424–6.203; p = 0.004) using univariate and 2.800 (95% CI, 1.335–5.873; p = 0.006) using multivariate analyses. The Borderline ASCVD group showed an elevated, though not statistically significant, risk using both univariate (HR, 1.533; 95% CI, 0.594–3.954; p = 0.377) and multivariate (aHR, 1.530; 95% CI, 0.593–3.950; p = 0.379).

In regard to the assessment based on CAD-RADS categories alone, higher CAD-RADS categories were associated with incrementally elevated HR for 5-year MACE compared with CAD-RADS 0 group using both univariate and multivariate analyses. Using univariate analyses, CAD-RADS 3 had an HR of 4.026 (95% CI, 1.669–9.709; p = 0.002), and CAD-RADS 4-5 an HR of 4.646 (95% CI, 2.415–8.938; p < 0.001). Using multivariate analyses, CAD-RADS 3 had an aHR of 3.897 (95% CI, 1.610–9.434; p = 0.003), and CAD-RADS 4-5 an aHR of 4.490 (95% CI, 2.329–8.655; p < 0.001). The CAD-RADS 1-2 showed an elevated, though not statistically significant, risk using both univariate (HR, 1.693; 95% CI, 0.931–3.082; p = 0.085) and multivariate (aHR, 1.616; 95% CI, 0.885–2.951; p = 0.119). These findings suggest that the anatomical extent and severity of CAD, as classified by CAD-RADS, are independent predictors of 5-year MACE in this outpatient cohort.

When CCTA findings were integrated with ASCVD risk categories, a more refined risk stratification for MACE was observed. Using the “ASCVD < 7.5% without OCAD” group as the reference, the presence of OCAD significantly increased the hazard for MACE within both dichotomized ASCVD risk strata. Patients with “ASCVD < 7.5% with OCAD” had a significantly elevated risk of MACE using both univariate (HR, 3.701; 95% CI, 1.218–11.244; p = 0.021) and multivariate analyses (aHR, 3.634; 95% CI, 1.194–11.059; p = 0.023). Patients with “ASCVD ≥ 7.5% without OCAD” showed a significantly increased risk of MACE using univariate analysis (HR, 1.921; 95% CI, 1.025–3.600; p = 0.041), but not using multivariate analyses (aHR, 1.840; 95% CI; 0.979–3.458; p = 0.058). The highest hazard for MACE was observed in patients with “ASCVD ≥ 7.5% with OCAD”, demonstrating robust significance using both univariate (HR, 5.294; 95% CI, 2.709–10.347; p < 0.001) and multivariate analyses (aHR, 5.101; 95% CI, 2.606–9.988; p < 0.001). Regarding sensitivity analyses using 5-year MACE-UCR as the outcome (Table S1), the results were similar to those of the primary analyses except that “ASCVD < 7.5% with OCAD” had no significant differences in 5-year MACE-UCR using both univariate and multivariate analyses.

## Discussion

CVD remains a major global health concern, highlighting the critical need for reliable risk stratification tools for guiding clinical decisions and improve patient outcomes. This study aimed to evaluate the diagnostic and prognostic utility of CCTA in outpatients presenting with stable chest pain, particularly in relation to the established 10-year ASCVD risk score. We identified cardiovascular risk factors through history-taking, chest radiography, electrocardiography, and basic blood examinations, then categorized patients by 10-year ASCVD risk scores and the presence of OCAD based on CCTA findings. CCTA, widely recognized for its clinical usefulness^15,18,19–22^ and recommended by the European and American guidelines for CAD diagnosis in stable chest pain patients^5,16,23,24^, allowed us to anatomically confirm and classify the degree of coronary stenosis using CAD-RADS categories. Adding CCTA findings into 10-year ASCVD risk score made MACE prediction more accurate, particularly in patients with ASCVD < 7.5% whose anatomic burden could be underestimated.

### Prevalence of OCAD in the Cohort Population

In our study, the prevalence of OCAD confirmed by CCTA among patients visiting an outpatient cardiology clinic with stable chest pain in this cohort was 16.0%. To our best knowledge, this is the first study in a Korean outpatient chest pain cohort to demonstrate that CCTA, when integrated with ASCVD risk, enhances MACE prediction over a 5-year period. Although a previous cross-sectional study in Korea analyzed 601 healthy health check-up examinees without known CVD, reporting a CCTA-confirmed coronary stenosis prevalence of 28.8%^25^, our investigation focused on symptomatic patients. Thus, our findings are crucial as they endorse the usefulness of CCTA as a recommended first-line diagnostic tool for stable chest pain in the Korean population, especially following recent guideline updates^5,17^. Our study also revealed a progressive increase in OCAD prevalence across higher ASCVD risk categories, which aligns with the established understanding that conventional risk factors contribute to the development of atherosclerosis.

### Comparison with Preceding Trials

Our findings align with and further extend the insights from landmark trials such as the PROMISE and SCOT-HEART regarding the diagnostic and prognostic utility of CCTA in patients with stable chest pain. The PROMISE trial demonstrated that a CCTA-first strategy was non-inferior to functional testing in terms of clinical outcomes^15^, while the SCOT-HEART trial notably showed that CCTA, when added to standard care, led to a reduction in non-fatal myocardial infarction and cardiovascular death by improving diagnostic accuracy and guiding management^18^. Consistent with these trials, our study reaffirms CCTA’s robust ability to detect and characterize CAD.

Our research uniquely contributes by specifically evaluating the additional prognostic value of CCTA findings beyond established ASCVD risk scores in a real-world outpatient setting. Although the PROMISE and SCOT-HEART established CCTA’s role in guiding overall patient management and improving outcomes compared with traditional pathways, the SCOT-HEART 10-year outcome analysis also suggested that, despite not reaching statistical significance, the point estimates for benefit from CCTA were generally greater in lower-risk groups, likely due to the identification of previously undetected coronary disease^26^. Building upon this observation, our study directly addresses a critical gap by demonstrating how CCTA can refine risk stratification and uncover significant anatomical disease even in individuals categorized as low or borderline risk by conventional 10-year ASCVD risk scores. This highlights CCTA’s potential not only as a diagnostic tool but also as a crucial adjunct for individualized risk prediction, ensuring that high-risk patients are not overlooked by traditional models.

### Clinical Implications of CCTA Integration

In our analysis of ROC curves, a clear hierarchy of diagnostic accuracy for detecting OCAD was observed, with CACS demonstrating significantly superior performance compared to symptom-based (DF) and traditional risk scores (FRS, ASCVD). This finding highlights a critical limitation of relying solely on subjective complaints of chest pain or clinical models, which often fail to precisely reflect the underlying anatomical burden of CAD.

Our study highlights the significant incremental prognostic value of CCTA beyond traditional ASCVD risk scores. By integrating the presence of OCAD, CCTA enhances risk stratification and improves prognostic accuracy across all ASCVD risk strata. Notably, we observed that a significant subset of patients with ASCVD < 7.5% demonstrated anatomically significant disease, as evidenced by a substantial incidence of MACE in “ ASCVD < 7.5% with OCAD” group compared with “ASCVD < 7.5% without OCAD” group. This reflects a critical discordance between calculated clinical risk and the actual anatomical burden of disease, demonstrating that anatomical evidence of disease via CCTA provides powerful prognostic insight, even in those deemed low risk by conventional ASCVD risk score. A recent study has reported a 2.7-fold higher risk of events in patients with ASCVD < 7.5% with OCAD compared with those with ASCVD ≥ 7.5% without OCAD^27^, a finding that our results further support.

These findings have substantial implications for patient care, specifically in the outpatient evaluation of stable chest pain. The detection of OCAD among individuals with low or borderline ASCVD risk reveals a hidden high-risk population that might otherwise be reassured by traditional scores alone. This mismatch can lead to delayed or missed opportunities for appropriate preventive interventions. Our data supports the integration of CCTA as a critical adjunct to existing risk models, particularly in symptomatic patients with inconclusive or low clinical risk profiles.

Our findings suggest that routine use of CCTA may be beneficial in selected outpatient populations, including those falling into lower ASCVD risk categories. Although the current guidelines endorse CCTA for selecting cases^5,16,23,24^, our results suggest a broader utility: In symptomatic patients with borderline ASCVD risk, identifying OCAD could prompt initiation of statin therapy, aspirin, or lifestyle counseling, ultimately improving long-term cardiovascular outcomes. This strategy reflects a more proactive, imaging-guided approach to cardiovascular risk management in the ambulatory setting.

### Strengths and Limitations

Our study benefits from its prospective, real-world clinical practice design and a relatively large cohort of 1,492 patients followed for up to 5 years, providing reliable insights into the long-term prognostic value of CCTA.

However, certain limitations should be acknowledged. First, as a single-center study conducted in Jecheon, Korea, its findings may not fully represent the general population, and generalizability to other populations warrants further investigation through multicenter studies. In addition, as an open-label, prospective study, the knowledge about the coronary CT results might have improved outcomes by inducing more aggressive treatment to patients. Second, the 10-year ASCVD risk score is primarily designed for 10-year outcomes in the general population, whereas our study evaluated 5-year MACE in a symptomatic cohort. Although ASCVD demonstrated strong predictive value in our symptomatic patients, this temporal difference should be considered. Third, although some variables were adjusted for multivariable analysis, 10-year ASCVD risk score and CCTA findings are correlated and unmeasured confounders might have still existed. Lastly, although our event rate was substantial (7.2% overall MACE), the relatively small number of overall events (n=108) limited our ability to perform more granular subgroup analyses with adequate statistical power, such as stratifying into more than two ASCVD risk groups when combined with CCTA findings.

Future research could explore the cost-effectiveness of routine CCTA in patients with lower ASCVD risk stable chest pain and investigate the impact of CCTA-guided management on actual MACE reduction in a randomized controlled trial setting.

## Conclusion

Our study demonstrates that CCTA-based detection of OCAD in symptomatic outpatients significantly improves 5-year MACE prediction, even among those classified as low-risk by 10-year ASCVD risk score, highlighting the clinical utility of anatomical imaging for risk stratification in ambulatory patients with stable chest pain. Incorporating CCTA into routine outpatient evaluation for stable chest pain can identify high-risk individuals who are underestimated by conventional clinical risk models. By facilitating earlier risk identification, this approach has the potential for improving preventive strategies and long-term outcomes, pending prospective validation.

## Data Availability

Data underlying this article will be available within the published article and from the corresponding author upon reasonable request.

## Acknowledgements

We would like to thank the radiologist Dr. Ik Soo Kim for the advice regarding cardiac CT measurement.

## Supplemental Material

Figure S1 Table S1

**Non-standard Abbreviations and Acronyms**
ACC/AHA: American College of Cardiology/American Heart Association
aHR: adjusted hazard ratio
ASCVD: atherosclerotic cardiovascular disease
AUC: area under the curve
CABG: coronary artery bypass grafting
CACS: coronary artery calcium scoring
CAD: coronary artery disease
CAD-RADS: Coronary Artery Disease-Reporting and Data System
CCTA: coronary computed tomography angiography
CHD: coronary heart disease
CKD: chronic kidney disease
CLEAR-OUT: Chest pain Longitudinal Evaluation with Advanced imaging for Risk stratification in an Outpatient cohort
CI: confidence interval
CT: computed tomography
DF: Diamond–Forrester
DM: diabetes mellitus
FRS: Framingham 10-year risk score
HR: hazard ratio
MACE: major adverse cardiovascular event
MACE-UCR: MACE excluding unplanned coronary revascularization
MI: myocardial infarction
OCAD: obstructive coronary artery disease
PCI: percutaneous coronary intervention
PROMISE: Prospective Multicenter Imaging Study for Evaluation of Chest Pain
ROC: receiver operating characteristic
SCOT-HEART: Scottish Computer Tomography of the Heart Trial
UA: unstable angina

## References

1. Abubakar I, Tillmann T, Banerjee A. Global, regional, and national age-sex specific all-cause and cause-specific mortality for 240 causes of death, 1990-2013: a systematic analysis for the Global Burden of Disease Study 2013. Lancet 2015;385:117–171.

2. Diamond GA, Forrester JS. Analysis of probability as an aid in the clinical diagnosis of coronary-artery disease. N. Engl. J. Med. 1979;300:1350–1358.

3. Kannel WB, McGee D, Gordon T. A general cardiovascular risk profile: the Framingham Study. Am. J. Cardiol. 1976;38:46–51.

4. Stone NJ, Robinson JG, Lichtenstein AH, Bairey Merz CN, Blum CB, Eckel RH, Goldberg AC, Gordon D, Levy D, Lloyd-Jones DM, McBride P, Schwartz JS, Shero ST, Smith SC, Watson K, Wilson PWF, American College of Cardiology/American Heart Association Task Force on Practice Guidelines. 2013 ACC/AHA guideline on the treatment of blood cholesterol to reduce atherosclerotic cardiovascular risk in adults: a report of the American College of Cardiology/American Heart Association Task Force on Practice Guidelines. J. Am. Coll. Cardiol. 2014;63:2889–2934.

5. Gulati M, Levy PD, Mukherjee D, et al. AHA/ACC/ASE/CHEST/SAEM/SCCT/SCMR guideline for the evaluation and diagnosis of chest pain: A report of the American College of Cardiology/American Heart Association joint committee on clinical practice guidelines. Circulation 2021;144:e368–e454.

6. Nilsson S, Scheike M, Engblom D, Karlsson LG, Mölstad S, Akerlind I, Ortoft K, Nylander E. Chest pain and ischaemic heart disease in primary care. Br. J. Gen. Pract. 2003;53:378–382.

7. Frese T, Mahlmeister J, Heitzer M, Sandholzer H. Chest pain in general practice: frequency, management, and results of encounter. J. Fam. Med. Prim. Care 2016;5:61–66.

8. Haasenritter J, Biroga T, Keunecke C, Becker A, Donner-Banzhoff N, Dornieden K, Stadje R, Viniol A, Bösner S. Causes of chest pain in primary care--a systematic review and meta-analysis. Croat. Med. J. 2015;56:422–430.

9. Wasfy MM, Brady TJ, Abbara S, Nasir K, Ghoshhajra BB, Truong QA, Hoffmann U, Di Carli MF, Blankstein R. Comparison of the Diamond-Forrester method and Duke Clinical Score to predict obstructive coronary artery disease by computed tomographic angiography. Am. J. Cardiol. 2012;109:998–1004.

10. Jang JJ, Krishnaswami A, Hung Y-Y. Predictive values of Framingham risk and coronary artery calcium scores in the detection of obstructive CAD in patients with normal SPECT. Angiology 2012;63:275–281.

11. Patel MR, Peterson ED, Dai D, Brennan JM, Redberg RF, Anderson HV, Brindis RG, Douglas PS. Low diagnostic yield of elective coronary angiography. N. Engl. J. Med. 2010;362:886–895.

12. Maffei E, Palumbo A, Martini C, Meijboom W, Tedeschi C, Spagnolo P, Zuccarelli A, Weustink A, Torri T, Mollet N, Seitun S, Krestin GP, Cademartiri F. Diagnostic accuracy of 64-slice computed tomography coronary angiography in a large population of patients without revascularisation: registry data and review of multicentre trials. Radiol. Med. 2010;115:368–384.

13. Seitun S, Clemente A, Maffei E, Toia P, La Grutta L, Cademartiri F. Prognostic value of cardiac CT. Radiol. Med. 2020;125:1135–1147.

14. Smulders MW, Jaarsma C, Nelemans PJ, Bekkers SCAM, Bucerius J, Leiner T, Crijns HJGM, Wildberger JE, Schalla S. Comparison of the prognostic value of negative non-invasive cardiac investigations in patients with suspected or known coronary artery disease–a meta-analysis. Eur. Heart J. Cardiovasc. Imaging 2017;18:980–987.

15. Douglas PS, Hoffmann U, Patel MR, Mark DB, Al-Khalidi HR, Cavanaugh B, Cole J, Dolor RJ, Fordyce CB, Huang M, Khan MA, Kosinski AS, Krucoff MW, Malhotra V, Picard MH, Udelson JE, Velazquez EJ, Yow E, Cooper LS, Lee KL, PROMISE Investigators. Outcomes of anatomical versus functional testing for coronary artery disease. N. Engl. J. Med. 2015;372:1291–1300.

16. Excellence NIfHaC. Chest pain of recent onset: assessment and diagnosis of recent onset chest pain or discomfort of suspected cardiac origin. London: national clinical guideline Centre for Acute and Chronic Conditions.; 2016. Accessed 2020 22 May. Available at http://www.nice.org.uk/guidance/CG95.

17. Cury RC, Leipsic J, Abbara S, Achenbach S, Berman D, Bittencourt M, Budoff M, Chinnaiyan K, Choi AD, Ghoshhajra B, Jacobs J, Koweek L, Lesser J, Maroules C, Rubin GD, Rybicki FJ, Shaw LJ, Williams MC, Williamson E, White CS, Villines TC, Blankstein R. CAD-RADS™ 2.0 – 2022 coronary artery disease-reporting and data system. J. Am. Coll. Cardiol. Imaging 2022;15:1974–2001.

18. CT coronary angiography in patients with suspected angina due to coronary heart disease (Scot-HEART): an open-label, parallel-group, multicentre trial. Lancet 2015;385:2383–2391.

19. Scot-HEART Investigators, Newby DE, Adamson PD, Berry C, Boon NA, Dweck MR, Flather M, Forbes J, Hunter A, Lewis S, MacLean S, Mills NL, Norrie J, Roditi G, Shah ASV, Timmis AD, van Beek EJR, Williams MC. Coronary CT angiography and 5-year risk of myocardial infarction. N. Engl. J. Med. 2018;379:924–933.

20. Chang H-J, Lin FY, Gebow D, An HY, Andreini D, Bathina R, Baggiano A, Beltrama V, Cerci R, Choi E-Y, Choi J-H, Choi S-Y, Chung N, Cole J, Doh J-H, Ha S-J, Her A-Y, Kepka C, Kim J-Y, Kim J-W, Kim S-W, Kim W, Pontone G, Valeti U, Villines TC, Lu Y, Kumar A, Cho I, Danad I, Han D, Heo R, Lee S-E, Lee JH, Park H-B, Sung J-M, Leflang D, Zullo J, Shaw LJ, Min JK. Selective referral using CCTA versus direct referral for individuals referred to invasive coronary angiography for suspected CAD: A randomized, controlled, open-label trial. JACC Cardiovasc. Imaging 2019;12:1303–1312.

21. DISCHARGE Trial Group, Maurovich-Horvat P, Bosserdt M, Kofoed KF, Rieckmann N, Benedek T, Donnelly P, Rodriguez-Palomares J, Erglis A, Štěchovský C, Šakalyte G, Čemerlić Adić N, Gutberlet M, Dodd JD, Diez I, Davis G, Zimmermann E, Kępka C, Vidakovic R, Francone M, Ilnicka-Suckiel M, Plank F, Knuuti J, Faria R, Schröder S, Berry C, Saba L, Ruzsics B, Kubiak C, Gutierrez-Ibarluzea I, Schultz Hansen K, Müller-Nordhorn J, Merkely B, Knudsen AD, Benedek I, Orr C, Xavier Valente F, Zvaigzne L, Suchánek V, Zajančkauskiene L, Adić F, Woinke M, Hensey M, Lecumberri I, Thwaite E, Laule M, Kruk M, Neskovic AN, Mancone M, Kuśmierz D, Feuchtner G, Pietilä M, Gama Ribeiro V, Drosch T, Delles C, Matta G, Fisher M, Szilveszter B, Larsen L, Ratiu M, Kelly S, Garcia Del Blanco B, Rubio A, Drobni ZD, Jurlander B, Rodean I, Regan S, Cuéllar Calabria H, Boussoussou M, Engstrøm T, Hodas R, Napp AE, Haase R, Feger S, Serna-Higuita LM, Neumann K, Dreger H, Rief M, Wieske V, Estrella M, Martus P, Dewey M. CT or invasive coronary angiography in stable chest pain. N. Engl. J. Med. 2022;386:1591–1602.

22. Machado MF, Felix N, Melo PHC, Gauza MM, Calomeni P, Generoso G, Khatri S, Altmayer S, Blankstein R, Bittencourt MS, Cardoso R. Coronary computed tomography angiography versus invasive coronary angiography in stable chest pain: A meta-analysis of randomized controlled trials. Circ. Cardiovasc. Imaging 2023;16:e015800.

23. Vrints C, Andreotti F, Koskinas KC, Rossello X, Adamo M, Ainslie J, Banning AP, Budaj A, Buechel RR, Chiariello GA, Chieffo A, Christodorescu RM, Deaton C, Doenst T, Jones HW, Kunadian V, Mehilli J, Milojevic M, Piek JJ, Pugliese F, Rubboli A, Semb AG, Senior R, Ten Berg JM, Van Belle E, Van Craenenbroeck EM, Vidal-Perez R, Winther S, ESC Scientific Document Group. 2024 ESC Guidelines for the management of chronic coronary syndromes: developed by the task force for the management of chronic coronary syndromes of the European Society of Cardiology (ESC) Endorsed by the European Association for Cardio-Thoracic Surgery (EACTS). Eur. Heart J. 2024;45:3415–3537.

24. Moss AJ, Williams MC, Newby DE, Nicol ED. The updated NICE guidelines: cardiac CT as the first-line test for coronary artery disease. Curr. Cardiovasc. Imaging Rep. 2017;10:15.

25. Noh DW, Kim S. Associations between coronary artery stenosis detected by coronary computed tomography angiography and the characteristics of health checkup examinees in the Republic of Korea. Radiography (Lond) 2020;26:22–26.

26. Williams MC, Wereski R, Tuck C, Adamson PD, Shah ASV, van Beek EJR, Roditi G, Berry C, Boon N, Flather M, Lewis S, Norrie J, Timmis AD, Mills NL, Dweck MR, Newby DE, Scot-HEART Investigators. Coronary CT angiography-guided management of patients with stable chest pain: 10-year outcomes from the Scot-HEART randomised controlled trial in Scotland. Lancet 2025;405:329–337.

27. Foldyna B, Mayrhofer T, Lu MT, Karády J, Kolossváry M, Ferencik M, Shah SH, Pagidipati NJ, Douglas PS, Hoffmann U. Prognostic value of CT-derived coronary artery disease characteristics varies by ASCVD risk: insights from the PROMISE trial. Eur. Radiol. 2023;33:4657–4667.

